# Empirical Review of LLM-driven Classification of Multidimensional Sleep Health Mentions from Free-Text Clinical Notes

**DOI:** 10.1101/2025.06.04.25328983

**Authors:** Syed-Amad Hussain, Ariana Calloway, Joseph Sirrianni, Eric Fosler-Lussier, Mattina Davenport

## Abstract

Accurate multidimensional sleep health (MSH) information is often fragmented and inconsistently represented within hospital infrastructures, leaving crucial details buried in unstructured clinical notes rather than discrete fields. This inconsistency complicates large-scale phenotyping, secondary analyses, and clinical decision support regarding sleep-related outcomes. In this work, we systematically explore contemporary natural language processing techniques, prompt-based large language models (LLMs) and fine-tuned discriminative classifiers, to bridge this critical gap. We evaluate performance on extracting nine key MSH dimensions (timing, duration, efficiency, sleep disorders, daytime sleepiness, interventions, medication, behavior, and satisfaction) from clinical narratives using public datasets (MIMIC-III derivatives) and an internally annotated pediatric sleep corpus.

Initially, we assess generative LLM performance using dynamic few-shot prompting, analyzing impacts from varying prompt structures, example quantity, and domain-specificity without explicit task-specific fine-tuning. Subsequently, we fine-tune generative LLM architectures on both in-task and out-of-task data to quantify performance improvements and limitations. Lastly, we benchmark these generative approaches against encoder-based discriminative classifiers (ModernBERT), designed to directly estimate binary presence of each MSH class within full clinical notes.

Our experiments demonstrate that fine-tuned discriminative models consistently provide higher classification accuracy, lower inference latency, and more robust span-level identification than either prompted or fine-tuned generative LLMs, given adequate training data. Nonetheless, generative LLMs retain moderate utility in low-data scenarios. Importantly, our results highlight persistent challenges, including difficulty extracting subtle sleep constructs such as sleep efficiency and daytime sleepiness, and biases associated with patient demographics and clinical departments. We conclude by suggesting future research directions: refining span extraction methods, mitigating biases in model performance, and exploring advanced chain-of-thought prompting techniques to achieve reliable, scalable MSH phenotyping within real-world clinical systems.

## Introduction

Accurate extraction of multidimensional sleep health (MSH) information from clinical narratives presents a critical opportunity for enhancing patient care, epidemiological research, and healthcare resource management. Clinical notes, often unstructured and narrative in nature, contain rich descriptions of patient behaviors, treatment interventions, and clinician observations, making them valuable but challenging sources for extracting nuanced clinical concepts such as sleep health. Traditional approaches to clinical natural language processing (NLP) have predominantly leveraged rule-based or simple machine learning models, often optimized for specific, narrow tasks and reliant on manually curated vocabularies or regular expressions. While such methods have shown high performance in limited scenarios, their ability to generalize across diverse clinical contexts and subcategories is often limited.

Recent advancements in large language models (LLMs), pre-trained on vast textual corpora, suggest significant potential to overcome these limitations through their enhanced semantic understanding and generalization capabilities. Domain-specific and clinically pretrained models, such as BioClinicalBERT and MentalBERT, have demonstrated superior performance over generic models by capturing nuances unique to medical contexts, including specialized terminologies and clinical reasoning. Moreover, tasks involving binary or multi-label classification approaches have shown varied effectiveness depending on their complexity and domain, highlighting the importance of selecting appropriate model architectures and training paradigms.

In this work, we systematically evaluate the effectiveness of contemporary LLMs, both generative (decoder-only) and discriminative (encoder-only), in identifying nine distinct categories of sleep-related information from clinical notes: behaviors, satisfaction/quality, daytime sleepiness/alertness, timing, efficiency, duration, disorders, medication, and interventions. Specifically, we investigate three core research questions: (1) How effectively do generative LLMs perform using dynamic prompting approaches without explicit fine-tuning, and what factors (e.g., prompt structure, few-shot example quantity, and domain-specificity) influence their performance? (2) How does targeted fine-tuning of generative LLMs on sleep-specific data influence performance and reliability in clinical settings? (3) To what extent can discriminative encoder models, specifically fine-tuned for binary classification, outperform generative models, particularly in terms of accuracy, computational efficiency, and practical deployability?

Our comprehensive experiments utilize both publicly available clinical datasets (MIMIC-III derivatives: MDACE and SDOH) and an internally annotated dataset from Nationwide Children’s Hospital (NCH-Sleep). We explore model performance across different experimental configurations—including variations in context window size, few-shot prompting scenarios, domain-specific pretraining, and fine-tuning strategies—to thoroughly characterize performance limits and practical applicability. We additionally assess biases inherent in these methods by stratifying results according to patient demographics, departmental contexts, and MSH classes, providing critical insights into potential disparities in model performance.

We demonstrate that fine-tuned discriminative models significantly outperform prompted and fine-tuned generative models, achieving superior classification accuracy, computational efficiency, and clinical applicability, particularly when trained with a combination of related-domain and task-specific data. Conversely, generative LLMs are shown to provide moderate performance with extensive prompting, highlighting their utility primarily when no labeled training data is available. Our findings underscore the value of careful selection of model type, training strategy, and data sources, ultimately guiding practitioners toward optimal approaches for clinical NLP tasks focused on sleep health phenotyping.

## Background

Recently, natural language processing (NLP), particularly leveraging Large Language Models (LLMs), has gained attention for efficiently extracting multidimensional sleep health (MSH) information from clinical narratives. NLP methods have historically employed rule-based or keyword-driven strategies for clinical data extraction. For example, Horner et al. (2022) developed a rule-based algorithm to detect sleep complaints from primary care notes, achieving high specificity (91%) but limited sensitivity (53%), relying mainly on keyword patterns and regular expressions [14].

Expanding these methods, Siviarajkumar et al. (2024) systematically compared rule-based, traditional machine learning, and LLM-based approaches, demonstrating the superior performance of their enhanced keyword-driven rule-based strategy across sleep-related concepts (F1-scores ranging from 0.91 to 1.0) [15]. Furthermore, Sirrianni et al. (2025) refined sleep-related vocabulary sets with structured clinical terminologies and semantic embeddings, significantly improving recall (0.992 vs. 0.857) in identifying notes with sleep mentions, although their method was limited to note-level classification without identifying precise textual locations or differentiating subcategories [17].

In parallel areas, such as suicide phenotyping, recent research further underscores the importance of domain-specific pretraining. Li et al. (2024) employed various BERT-based models, including generic models like BERT and RoBERTa, domain-adapted models such as BioClinicalBERT, and disease-specific models like MentalBERT, specifically pre-trained on mental health text [16]. They demonstrated that domain-adapted and disease-specific models significantly outperformed generic models, highlighting the advantages of domain-specific pretraining for clinical NLP tasks [16]. Their binary classification strategy, transforming multi-label classification into multiple independent binary tasks, proved effective. However, they also found enhanced performance when employing single multi-label classifiers that could exploit label interdependencies [16].

These findings collectively indicate that while foundational NLP methods have effectively initiated sleep information extraction from clinical notes, advanced LLM architectures, particularly those with domain-specific pretraining and sophisticated task structures, are increasingly necessary. Such methods promise to better capture complex semantics and contextual subtleties inherent to clinical documentation related to multidimensional sleep health concepts.

## Data

Our work employs both de-identified MIMIC-III datasets (consisting of MDACE and SDOH as described in the MIMIC-III datasets section) as well as an internal sleep mentions dataset [1, 2]. All datasets, derived from electronic medical records, consist of notes which are annotated according to if a given class is present in the note. The quantity of notes, note sizes, total classes, and positive classes per note are described in Table 1.

**Table 1:** Basic metrics of datasets used. Note length reported in characters. Mean class refers to average number of positive classes per note.

| Dataset | # Notes | # Classes | Mean note length (std) | Mean class/note (std) |
| --- | --- | --- | --- | --- |
| MDACE [2] | 1192 | 1462 | 6380 (5507) | 6.3 (6.2) |
| SDOH [1] | 145 | 20 | 3103 (1772) | 1.3 (1.3) |
| NCH-Sleep | 300 | 9 | 6815 (5863) | 2.5 (1.9) |

## MIMIC-III Datasets

MIMIC-III (Medical Information Mart for Intensive Care) is a publicly available dataset of deidentified electronic health records for patients admitted to critical care at a large tertiary care hospital. While MIMIC-III includes many data points (e.g. laboratory measurements and medications), we are primarily focused on downstream datasets which make use of the free-text clinical notes within MIMIC-III [3]. These clinical note datasets serve as a proxy for our sleep mention classification task of positively identifying mentions of a given property within the free-text clinical note.

**MDACE:** In “MIMIC Documents Annotated with Code Evidence”, Cheng et al. (2023) provide a dataset with expert annotations over 302 inpatient charts [1]. The experts use ICD-9 and ICD-10 schemas to determine coding— we choose to focus on ICD-10 due to increased contemporary relevance [1]. Each of these annotated codes has an associated evidence span within the clinical note text, resulting in a total of 5,563 total evidence spans for the dataset [1]. Additionally, we gather class descriptions by cross referencing each annotated ICD-10 code with its definition as per CDC.gov.

**SDOH:** In “Annotation dataset of social determinants of health from MIMIC-III Clinical Care Database”, Guevera et al. (2024) annotate MIMIC-III clinical notes with patient status over 6 factors (e.g. employment, housing, transportation, parental status, relationship, and social support), totaling 20 possible statuses [2]. SDOH classification acts as a unique foil for our internal dataset as both have descriptive indicators primarily encoded within clinical text rather than in structured data fields. This work has a total of 145 expert annotated notes, which they expand using synthetic data generation [2]. We only make use of the expert annotated notes to avoid any noise generated by synthetic data points. Like MDACE, the annotations include evidence spans to justify the classification [1, 2].

### NCH-Sleep Dataset

We construct a sleep dataset, NCH-Sleep, using 300 clinical notes written between 2018 and 2023 at Nationwide Children’s Hospital (NCH). These clinical notes were initially pulled from the hospital EHR system using keyword matching over a list of expert-defined sleep-related keywords from four departments: primary care, behavioral health, healthy weight, and school based. We randomly sampled 300 notes from the entire dataset (∼700,000 notes) stratified by department and year (we additionally stratify our train/test split on departments as their note-taking structures have dramatically distinct styles). Subsequently, each note was expert annotated for presence of the 9 sleep MSH classes, with corresponding evidence spans highlighted. Table 2 describes the 9 sleep MSH classes and their relevant descriptions.Table 3 lists each department and their relative frequency of notes.

**Table 2:** The 9 MSH classes annotated in the NCH-Sleep dataset and evaluated in each of our experiments. Class descriptions are used to help inform models.

| MSH Class Name | Class Description | # Positive Notes in NCH-Sleep |
| --- | --- | --- |
| Timing | Mention of timing of sleep-related events | 40 / 300 |
| Sleep Disorder | Mention of a sleep disorder | 81 / 300 |
| Alertness and Daytime Sleepiness | Mention of alertness or daytime sleepiness | 77 / 300 |
| Behavior | Mention of sleep related behavior | 55 / 300 |
| Efficiency | Mention of sleep efficiency | 77 / 300 |
| Sleep Medication | Mention of sleep medication | 90 / 300 |
| Satisfaction | Mention of satisfaction with sleep | 64 / 300 |
| Duration | Mention of duration of sleep | 32 / 300 |
| Sleep Intervention | Mention of a sleep intervention | 51 / 300 |

**Table 3:** Departments and their respective number of notes within NCH-Sleep.

| Department | # Notes in NCH-Sleep |
| --- | --- |
| Primary Care | 174 |
| Behavioral Health | 106 |
| Healthy Weight | 14 |
| School Based | 6 |

For all analyses presented in this work we use a 67/33 (200 note / 100 note) train/test split over the NCH-Sleep dataset. Even when NCH-Sleep train data is not directly used for model training or context sampling, we still validate every method over the 100 note NCH-Sleep test set. We use this split ratio as opposed to the standard 80/20 due to the small size of our dataset and the need for a representative evaluation. Furthermore, while training tasks benefit from larger training sets, prompting methods do not share a similar scaling with training set size. This 67/33 split is meant to balance the needs for evaluation and training.

### Task Structure

#### Boolean Classification per Class

While our evaluation can be represented as a multi-class classification task over the 9 MSH classes, we assume conditional independence of each class, especially since multiple classes can be true at once for a given note. This assumption is beneficial as it allows us to frame our task as a boolean (true/false) classification for each given class, freeing evaluated models to focus on their prediction of one class at a time rather than allowing confusion between classes.

Additionally, since all the classes do not to be predefined, this Boolean construction allows for evaluated models to use examples from the MIMIC-III based datasets as an indicator of task structure, signaling to the model that classification must occur by attending class descriptions alongside a given clinical note. Finally, this construction has the benefit that a downstream model can be queried on a class unseen in training, allowing for clinicians to pivot the exact classes they are searching if the model proves good generalizability. The template construction for each example is as follows:

`Input: {‘clinical_note’: {note_text}, ‘class_label’: {class_name}, ‘class_description’: {class_description}}`

`Output: “True”/“False” for generative tasks; 1/0 for discriminative tasks`

#### Negative Sampling for Training Data

In each dataset included in this work, every note is annotated according to its positive classes. We assume that if a class present in the dataset is not part of the positive classes for a given note, it can be considered a negative class. Classification tasks are known to benefit from contrastive training in which negative examples are paired with positive examples, signaling to the model that it should shift the distribution of positive and negative examples away from each other.

This shift allows for better discrimination over a classification task.

We employ negative-sampling over each dataset to create our training sets, sampling 3 negative examples for every positive example. In NCH-Sleep and SDOH, the negative samples are randomly selected from the set of classes within the dataset not present among the positive classes for that note. In MDACE, we make use of the hierarchical structure of ICD-10 codes to sample negative classes at varying levels of similarity to the positive classes for a given note. We sample one negative class from the entire list of ICD-10 codes, one negative class that shares the first letter in its code with a code present in the note, and one negative class that shares the first three characters with a code present in the note. This staged sampling allows the training set to create varying levels of challenge for the model.

#### Evaluation

We conduct our evaluations over the 100-note test split of NCH-Sleep. To imitate a clinical use case, we evaluate every note for all 9 MSH classes rather than just the positive class and 3 negative samples as in the training set. We do, however, employ a version of this test split with 3 negative samples to use as a validation for fine-tuned models to determine the best training epoch for final evaluation.

## Experiment 1: Prompting LLMs

Contemporary LLMs benefit from extensive pre-training and high parameterization, allowing for in-context learning during inference [4, 5, 8]. This capability allows models to perform tasks without any explicit training over the target task. Performance can be adapted by changing the context provided to the model, for example, by manipulating prompt instructions, altering example context lengths, or providing task-representative examples. Models used without targeted training are pre-trained on corpuses which can be general or domain-specific (e.g. medical), potentially shifting their capabilities on a specific task. Such LLMs are generative, meaning they output sequences of text, however they calculate probabilities over each token in the process. In this section we will explore each of these properties as they relate to the classification of MSH over the NCH-Sleep test set.

**Runtime Parameters:** We conduct each experiment on an NVIDIA H100 with 80GB of VRAM and a batch size of 1 and int-4 quantization. Models are gathered from online repository Huggingface.

### Methods

**The Prompt:** We make use of the following prompt format:

`“””`

`You are a system that aids in information extraction from clinical notes.`

`You are provided a clinical note, a class name, and a class description.`

`You must classify if the class is present in the clinical note.`

`Your output must be only the word True or False.`

`Do not output anything other than the word True or False.`

`Use the following training examples to help refine your understanding of the task: {few_shot_examples}`

`Example to predict on: Input: {test_example} Output:`

`“””`

This format was determined through rapid evaluation using the MIMIC-III datasets over GPT-4o-mini (process and evaluation results described in Appendix A1) [8]. The line including ‘few_shot_examples’ is excluded when the number of few-shots to include is 0.

#### Dynamic Few-Shot Prompting

We select our few-shot examples through a dynamic method which seeks to identify the examples in a training set that are most similar to a given test example. Namely, we encode the input of each training example, as defined in the Task Definition, using an LM to create a pool of training embeddings. During inference we encode the input for the test example and find the closest *n* examples in the training embedding pool according to cosine distance, a standard metric of embedding similarity. These *n* examples are the ones selected to be the few-shot exemplars for the prompt, with *n* being an independent variable of our comparisons.

To generate the embeddings, we use ‘intfloat/e5-large-v2’, an encoder developed by Microsoft using weakly supervised contrastive pre-training [6]. This model was selected for its lightweight parameter size (560M) and strong encoder performance as ranked on the Huggingface MTEB (Massive Text Embedding Benchmark) leaderboard, allowing for fast and effective encoding [9]. Since the max context window of this model is only 512 tokens, which is much smaller than most of our example clinical notes, we perform encoding by sliding the input text in chunks of 512 with a stride of 256. We aggregate the final embedding by mean-pooling over these window encodings as described in Eq 1.

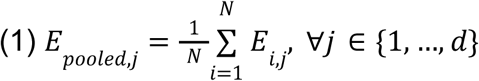

Where 𝑁 is the total number of windows, 𝐸_𝑖_ is the embedding vector for the 𝑖 𝑡ℎ window, 𝐸_𝑝𝑜𝑜𝑙𝑒𝑑_ is the final mean-pooled representation, and 𝑗 indexes the dimensions of the embeddings.

### Independent Variables

We control for a series of independent variables which are summarized in Table 4. Our evaluations compare each combination of these independent variables.

**Table 4:**
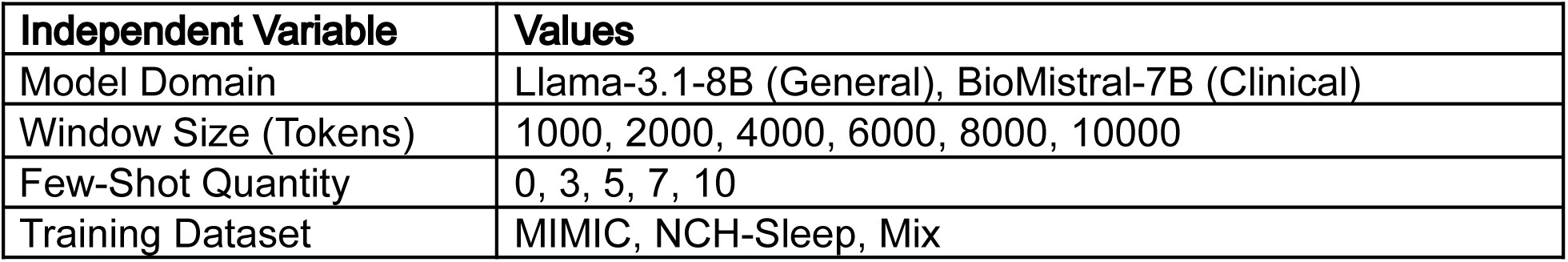
Independent variables and their relevant values tested using prompt-based LLM evaluation.

| Independent Variable | Values |
| --- | --- |
| Model Domain | Llama-3.1-8B (General), BioMistral-7B (Clinical) |
| Window Size (Tokens) | 1000, 2000, 4000, 6000, 8000, 10000 |
| Few-Shot Quantity | 0, 3, 5, 7, 10 |
| Training Dataset | MIMIC, NCH-Sleep, Mix |

#### Domain-specific vs General Domain Model

We compare two LLMs which, while having similar architectures, differ in their pretraining and subsequent instruction tuning. Namely we compare ‘meta-llama/Llama-3.1-8B’ and the slightly smaller ‘BioMistral/BioMistral-7B’, where 8B and 7B refer to their parameter count [4, 5]. Models of this parameter count were selected as they are the biggest that would fit on one 80gb GPU while maintaining high context-size. We select Llama-3.1-8B as our general model as it remains an industry standard benchmark for open-source LLMs [5]. Meanwhile, BioMistral-7B was selected because its pre-training on PubMed PMC oriented it towards clinical tasks while maintaining an otherwise similar architecture to Llama-3.1-8B [4, 5].

#### Window Size

While these models can handle a context length upwards of 8,000 tokens, many of our notes get close to or exceed this maximum value, especially when adding context needed by few-shot exemplars and the prompt itself. Compounding this issue, our 80Gb GPU is unable to handle the entirety of the 8,000 token context without running out of memory. As such, we instead chunk each note using sliding windows and perform classification on each chunk. This does however come with the drawback of limiting the information the model can make a prediction on. To understand the implications of context window sizes we evaluate performance over the set listed in Table 4.

For each window we have a stride that is window//2 (i.e. the window size of 1000 has a stride of 500), ensuring no subcomponent of the text is presented in an under-contextualized manner. For a given chunk, we define the classification only if the span indicating positive presence is entirely within the chunk. If the note has a positive classification but the chunk does not entirely contain the relevant span, the chunk is skipped when creating the training data as we cannot guarantee there is a positive or negative signal in the context of that chunk. If the note is negative for a given class, all chunks are included as negative examples for that class.

#### Few-Shot Example Quantity

Including examples of the task, with both inputs and expected outputs, helps to orient a model to the task. A general example can be selected to provide a high-level understanding of the task while more specifically chosen examples can be used to orient more nuanced semantic understandings of the model, as is the goal with the dynamic few-shot sampling method we employ here. It is unclear, however, how many examples are needed to assist the model, or what level performance increase these examples provide above just using the model’s pre-trained knowledge. To understand the implications of few-shot sample sizes we evaluate performance over the set listed in Table 4.

#### Few-shot Example Source (Training Dataset)

The source of the few-shot examples has the potential to have as much, if not more, effect than the quantity of examples provided. In some clinical tasks, often there is paucity-labelled data, necessitating the use of a distant signal from tangential datasets. We explore the effect of sample sources between in-task (NCH-Sleep), out-of-task (MIMIC), and a combination of the two (Mix). These comparisons allow us to understand how necessary annotated data is for validating MSH with clinical notes when using LLMs.

### Accessing the Predictions

#### Generative

LLMs are decoder-only models that autoregressively generate the next token after attending to a given input context [4, 5]. As such, they are trained to output tokens rather than classification probabilities. For our primary analyses with LLMs, we task them to generate the tokens “true” or “false” to classify MSH class presence. It is possible the LLM generates other tokens so we simply search the generation, which is limited to 10 tokens, and detect which ever token between “true” and “false” occurs first. If neither occurs, the window is skipped.

#### Token Probability

While LLMs are trained to output tokens, they decide this output by estimating a distribution over their vocabulary size (i.e. all the tokens it can consider) and sampling the most probable token [4]. This estimation is presented as a logit, a logarithm of the corresponding odds of a given token, as described in Eq. 2. To ensure we can always gather a prediction from the LLM even when its generation may go awry, we calculate a probability of the class being true directly using the logit values for the ‘true’ and ‘false’ token as it begins to generate its response, described in Eq. 3. It is, however, unclear how well these probabilities will map towards model confidence as this mechanism does not align with the LLM training objectives. If the mapping is effective, the probabilities can be used to create ROC curves that allow prediction systems to vary their thresholds to maximize clinically effective precision and recall.

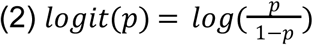

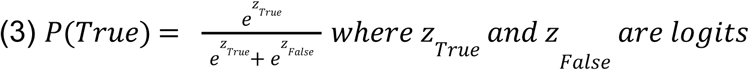

#### Note vs Window Level Prediction

To replicate the downstream clinical use case, we conduct most of our evaluations at the note level. If a given note is split into chunks due to windowing, we aggregate the prediction over each chunk so that if even one chunk is positive, the entire note is classified as positive. This method is necessary as it is possible that most of a note would not have the information needed for a classification save for the content in one chunk, therefore the one positive case needs to supersede all negative cases. When using token probability evaluation, we simply take the maximum probability over the set of chunks that constitute a note as our final prediction probability for the note.

We do, however, still present the window-level performance of our best performing system. This is because in clinical use case, if we employ a given model we can have the window level predictions help highlight chunks of the text that indicate positive presence of a given class. Given high window-level performance, this would help a clinician rapidly screen model decisions for veracity and bias. In practice, this would function as a second pass after a note had been initially classified.

### Results

We present a complete table of results for Experiment 1 in Appendix A2.

### Overall Results

Our top evaluated configuration was Llama-3.1-8B using a context window of 2000 characters and supplied with 10 few-shot exemplars, resulting in a precision of 0.25, recall of 0.83, and overall f1 score of 0.38. For all test configuration, the recall was higher than precision. This is an expected effect due to our evaluation technique which prioritizes a single positively classified window over all negatively classified windows for a given note. While high recall is preferred for screening tasks, the low overall F1, with little variation between all run configurations, implies that prompting LLMs is insufficient for clinical application of MSH classification.

Token Probability: Looking at “true” token probabilities directly, we find the effect of high-recall / low-precision is even stronger, with nearly all cases having a recall of 1 and therefore similar precisions of 0.21 as all notes incur as positive classification. We do see variance when looking at AUC however, finding the best performing AUC of 0.68 and the “top” system having an AUC of 0.64. This implies there may be a better cutoff for performance than 0.5 for the true probability, however the overall performance of the LLM is better suited for generative classification rather than by accessing token probabilities directly. This supports the hypothesis that generative systems do not learn class discrimination directly but rather fit to a token generation task.

Window-Level Performance: The top system had a precision of 0.36, recall of 0.31 and F1 score of 0.334 at the window level. This is not a high performance and, when paired with the middling performance at the note-level, indicates poor clinical applicability for informed decision making. Namely, the windows classifications are not likely informative for a clinician seeking to quickly scan a note for relevant text.

### Effect of Independent Variables

For each independent variable we compare the constituent values by gathering the average F1 score among all runs sharing that value. We present our results for the summarized effect of each independent variable in Table 5.

**Table 5:**
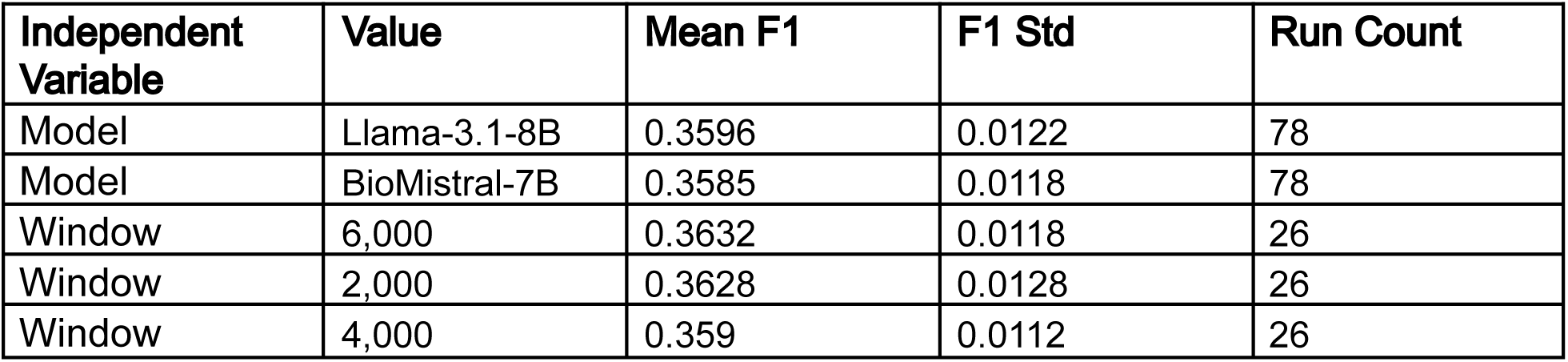
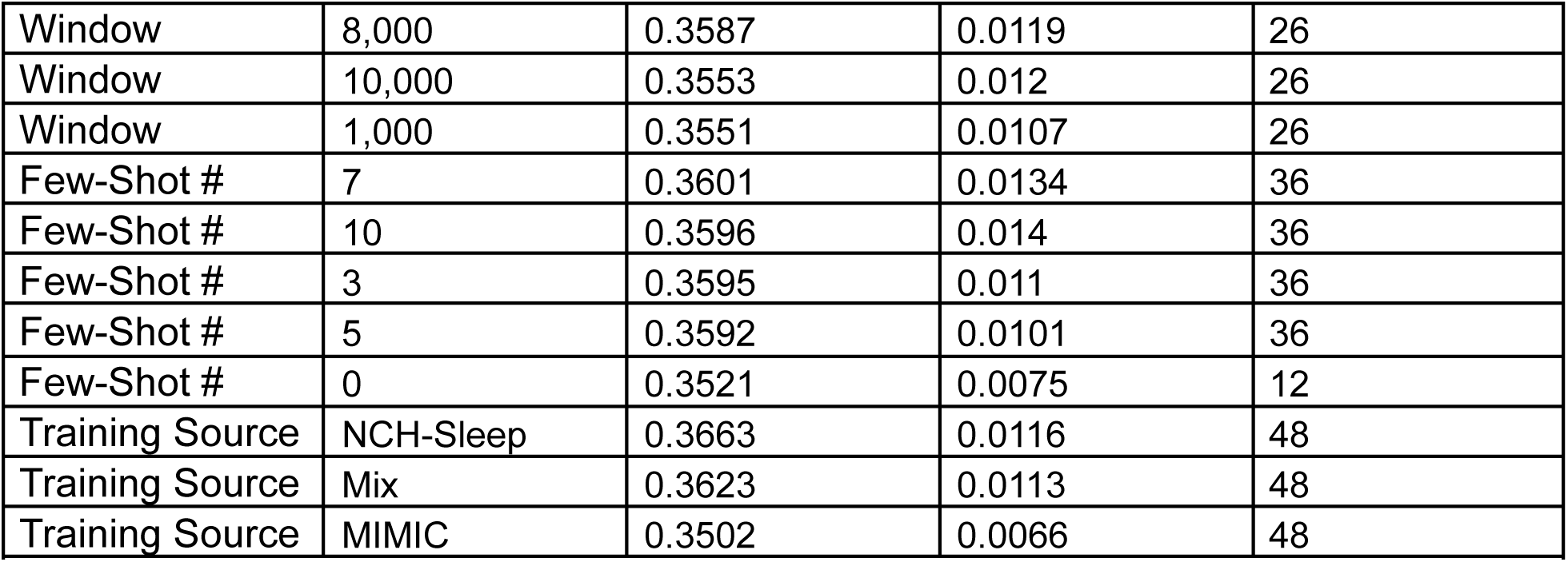
Comparison of the effect of each independent variable when prompting LLMs for classification. The results within each independent variable are sorted in descending order of F1 score.

**Model:** We see no substantial difference between our two model types despite BioMistral-7B being trained on biomedical literature (PubMed PMC) [4]. This lack of performance may indicate the task is challenging enough that neither model’s difference in capabilities are able to be apparent. Another reason may be that PubMed PMC, being medical literature, is not a good proxy for relatively unstructured clinical notes.

**Window:** We find little correlation between window size and performance; we cannot say higher window sizes incur better or worse performance. Rather we see the medial window sizes perform best, with 6000 as the top performer on average, while the most extreme values of 10,000 and 1,000 are the worst performers. This may be because the model has to balance having enough context to make an informed decision, while not using so much context that few-shot quantities must be lessened or context truncation must incur.

**Few Shot Quantity:** We find larger few-shot quantities correlate to better performance, with the 7 and 10 few-shot quantities performing best, albeit marginally. This may indicate that performance is best promoted when the model is sufficiently able to learn the task using few-shot exemplars. This finding is further supported by zero-shot performance, which proved the worst among the runs. All of our evidence thus indicates that model pre-training knowledge and prompt instructions alone are insufficient to properly encode the task.

**Training Data Source:** We find using NCH-Sleep data is the best informant for the task. This is to be expected as the training sample is meant to be representative of the examples found in the NCH-Sleep test set. Suprisingly, we find that Mix underperforms NCH-Sleep, indicating that providing MIMIC-examples can distract from good performance rather than aid it. This hypothesis is fortified when we see zero-shot performance outperforms MIMIC few-shot performance on average (F1 of 0.3521 vs F1 of 0.3502).

**Table 1a:**
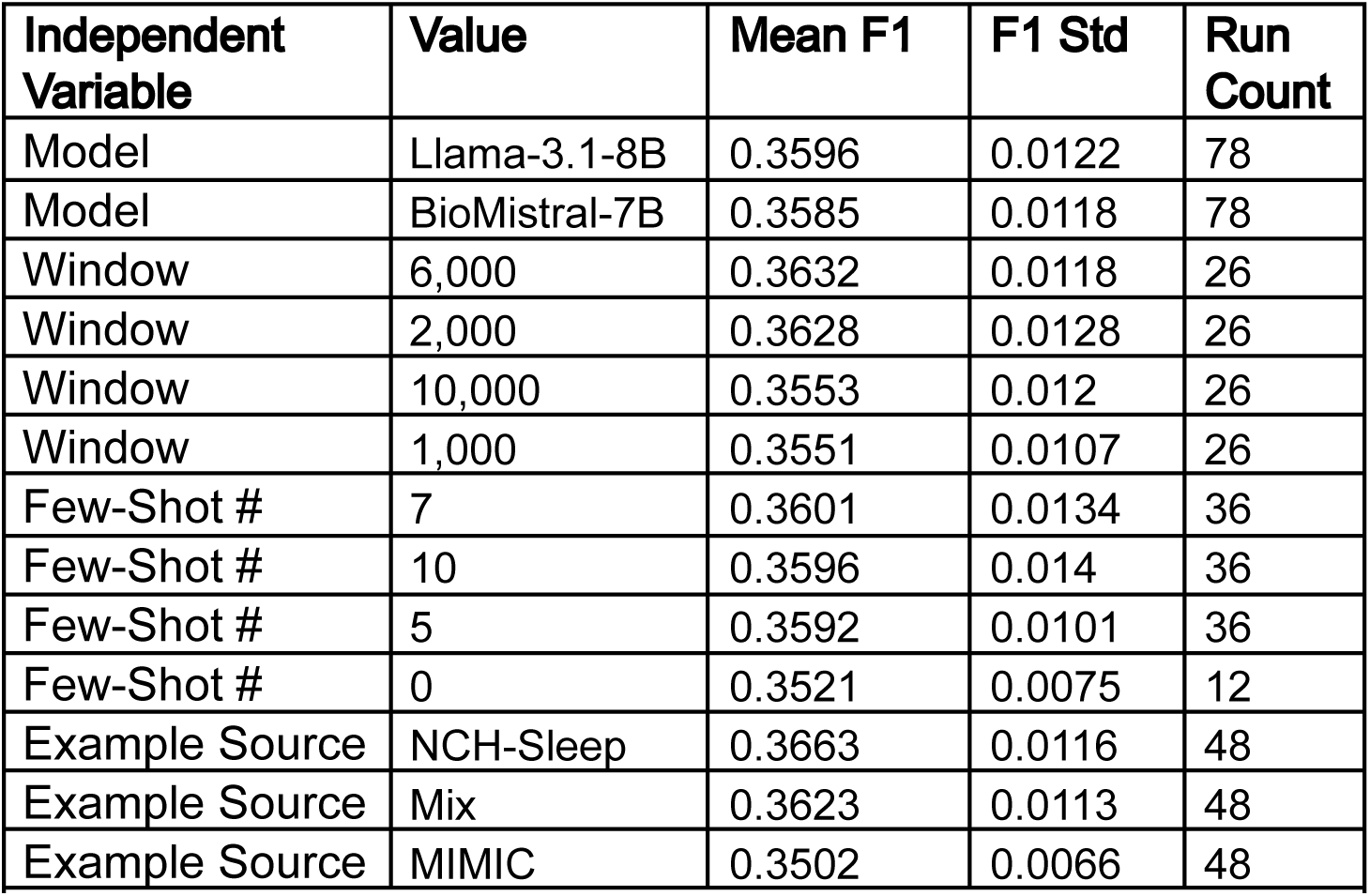
Comparison of the effect of select independent variables when prompting LLMs for classification. The results within each independent variable are sorted in descending order of F1 score.

## Experiment 2: Fine-tuned LLMs

The results from Experiment 1 show that higher few-shot counts correlate to better performance. This may indicate that one of the toughest challenges for the LLM is understanding the structure of the task itself. For this reason, fine-tuning of the LLM may be fruitful as this would allow the LLM to see every training example in our dataset and better orient towards the task.

### Methods

#### Model

We take the best performing model from Experiment 1, Llama-3.1-8B and fine-tune it over each training data source described in Table 4. A comparison over these training datasets allows us to understand the need and effect of having in-task data for training. We continue to evaluate on the 100 note NCH-Sleep test split.

#### Window Size

Experiment 1 results showed a minimal correlation between window size and performance; therefore we use the highest window size that reliably fits within the GPU VRAM limit of our NVIDIA H100, 8000 characters. We choose this value instead of the top performing value of 6000 as few-shot examples are not included during fine-tuning and therefore more context can be provided for the example to be tested.

#### Training Parameters

We train each model using an AdamW optimizer, cross entropy loss, and for up to 48 hours or 50 epochs, saving the epoch with the best performance over the validation set. Additionally, we use LORA, PEFT, and int-4 quantization for more efficient fine-tuning [10, 11].

### Results

We present results for our fine-tuned LLMs in Table 6. Once again, we find including information from NCH-Sleep in our training set to be effective, however the Mix dataset very slightly outperforms the NCH-Sleep dataset alone, potentially indicating some benefit to the added MIMIC examples. We do still find the NCH-Sleep trained model to have the best token probability AUC, implying better use in a risk stratification application.

**Table 6:** Results of Llama-3.1-8B fine-tuned over 3 training sets and evaluated on NCH-Sleep test.

| Training Data | Precision | Recall | F1 | Token Prob AUC | Skipped Notes | Skipped Windows |
| --- | --- | --- | --- | --- | --- | --- |
| Mix | 0.2912 | 0.5856 | 0.389 | 0.6245 | 35 | 231 |
| NCH-Sleep | 0.2663 | 0.7005 | 0.3859 | 0.6501 | 19 | 128 |
| MIMIC | 0.2811 | 0.522 | 0.3654 | 0.622 | 29 | 238 |

We additionally find the fine-tuned models had an increased risk of hallucinating invalid answers. In Table 6, we note “Skipped Windows” and “Skipped Notes” to show cases where the model did not generate “true” or “false”. The frequency of these failures increased from the frequency of failures by prompting methods (which was limited to a few windows and configurations). Hallucinations, which “Token Prob AUC” is robust to, shows the benefit of creating a discriminative prediction rather than a generative one.

Even so, when we mark these examples as incorrect, we find the F1 of each model is about 3 points better than the best prompting configuration, showing consistent— albeit small— gains over the prompting approach.

## Experiment 3: Fine-tuned Classifier

Generative LLM’s are trained to produce tokens rather than estimate classification probabilities directly. In this case, the generation of a “true” or “false” token only serves as a proxy for our task. Likewise, much of the parameterization of LLMs is to promote better generations and generalizability but may not be necessary for effective discrimination of classes, especially given availability of fine-tuning data.

In this experiment we fine-tune an encoder model to estimate classification probabilities directly. The benefits of building an effective encoder-based discriminative system include faster runtime, lower VRAM requirements allowing for larger context size, faster training convergence, and direct calculation of ROC for downstream risk stratification [7]. The potential downsides include worse generalizability and increased reliance on the quality of training data.

### Methods

#### Model

In this experiment we fine-tune ‘answerdotai/ModernBERT-basè, a bidirectional encoder that expands on the original BERT work through the implementation of contemporary model optimizations and training over 2 trillion tokens [7]. This model boasts memory and runtime efficiency alongside a sequence length limit of 8192 tokens [7].

#### Window Size

ModernBERT’s high token limit and VRAM efficiency allows us to avoid chunking notes into windows and rather classify each note in its entirety. Therefore we forgo the aggregation of chunk evaluations that leads to higher recall in our LLM based experiments.

#### Prediction Calculation

Rather than predict the token “true” or “false”, we encode each classification as the integer 1 (true) and 0 (false). The model predicts the probability of label 0 or 1. We report back the positive label probability by computing the softmax over these two-class probabilities as described in Eq. 4.

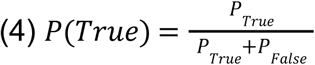

#### Training parameters

We train our models using NVIDIA A100 GPUs with 40GB of VRAM. We use the AdamW optimizer and binary cross entropy loss. We train for up to 200 epochs and 24 hours, selecting the best performing epoch over the 100 note

NCH-Sleep validation set

We again train over the 3 training datasets described in Table 4.

### Results

We present our results for our fine-tuned classifiers in Table 7. The order of training data effect in this work follows the results of our fine-tuned Llama-3.1-8B models in Experiment 2, with Mix data performing the best followed by NCH-Sleep and then MIMIC. The disparities in performance between these datasets are much larger however. We see the Mix trained model has the best F1 and AUC score of any system we have seen so far, while the NCH-Sleep trained model falls dramatically in performance by nearly 15 points F1 and 5 points AUC. Meanwhile the MIMIC trained model performs abysmally with an F1 of 0.07, a 35 point drop in performance from the best trained version.

**Table 7:** Results of ModernBERT fine-tuned over 3 training sets and evaluated on NCH-Sleep test.

| Training Data | Precision | Recall | F1 | AUC |
| --- | --- | --- | --- | --- |
| Mix | 0.4679 | 0.3923 | 0.4268 | 0.7083 |
| NCH-Sleep | 0.4804 | 0.1885 | 0.2707 | 0.6501 |
| MIMIC | 0.2075 | 0.0423 | 0.0703 | 0.4425 |

**Table 8:**
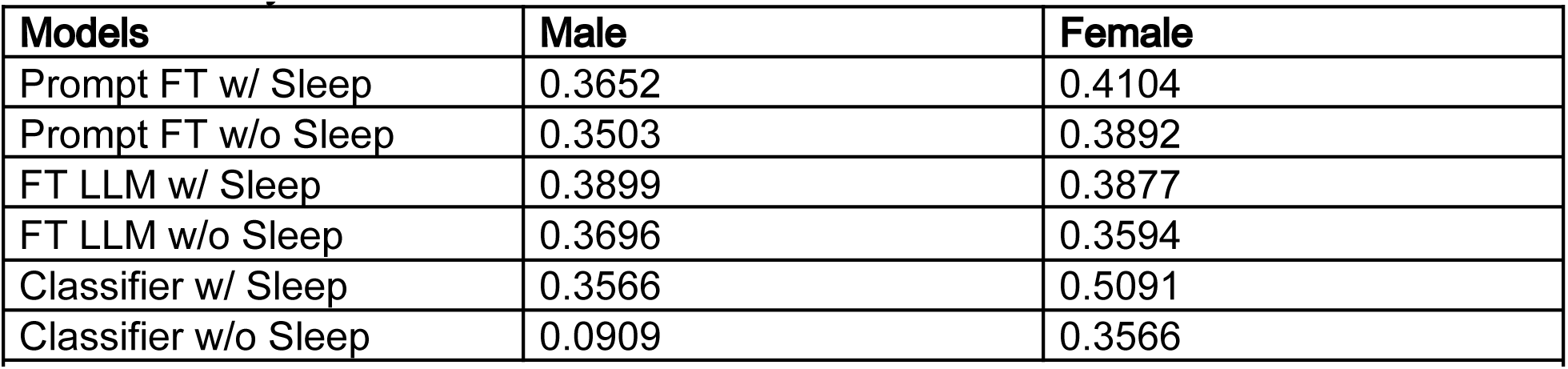
Performance (F1) of the top performing models, and their best non NCH-Sleep trained versions, when stratified by gender.

**Table 9:** Performance (F1) of the top performing models, and their best non NCH-Sleep trained versions, when stratified by race.

| Models | White | Latino | African American | Bi-Racial |
| --- | --- | --- | --- | --- |
| Prompt FT w/ Sleep | 0.4404 | 0.3438 | 0.3438 | 0.3810 |
| Prompt FT w/o Sleep | 0.4094 | 0.3803 | 0.3252 | 0.3448 |
| FT LLM w/ Sleep | 0.4164 | 0.3243 | 0.3759 | 0.4828 |
| FT LLM w/o Sleep | 0.3891 | 0.3030 | 0.3676 | 0.3846 |
| Classifier w/ Sleep | 0.5035 | 0.2540 | 0.3048 | 0.5833 |
| Classifier w/o Sleep | 0.0117 | 0.1277 | 0.1667 | 0.0000 |

**Table 10:**
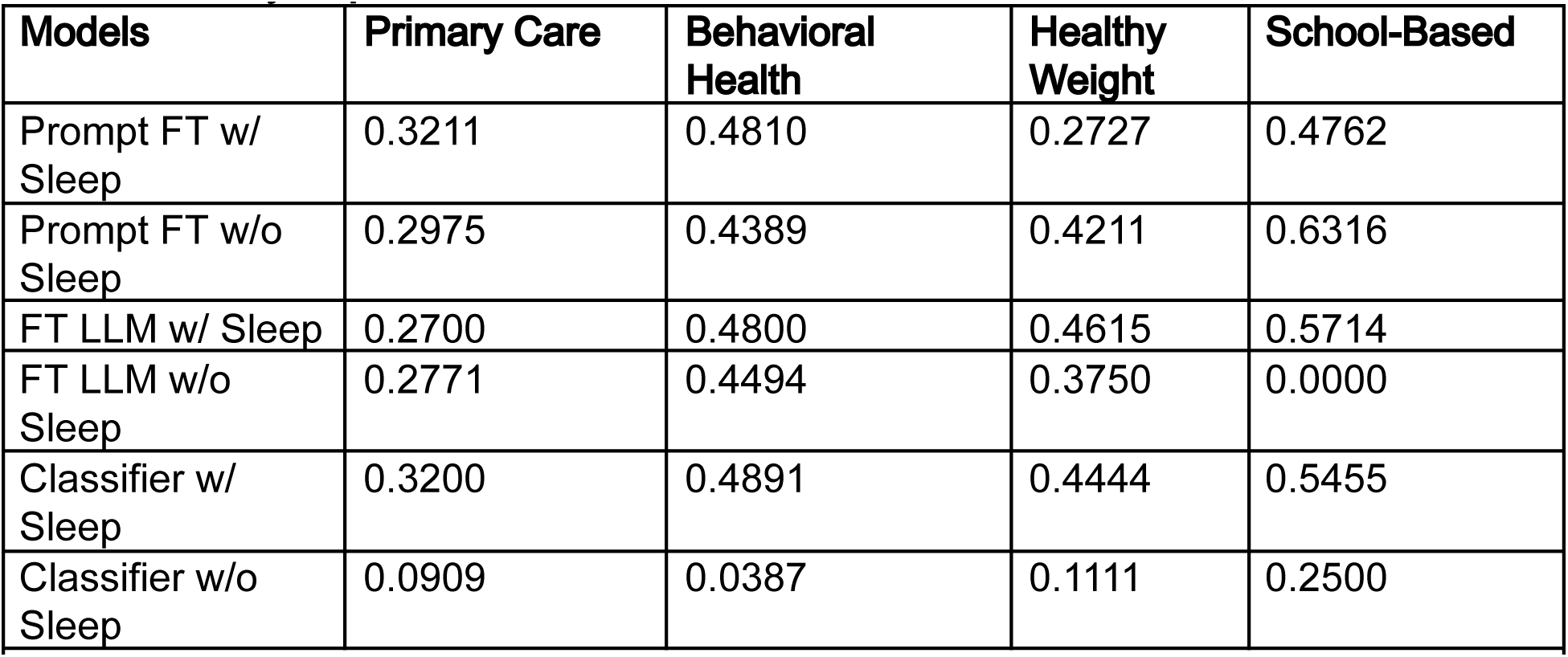
Performance (F1) of the top performing models, and their best non NCH-Sleep trained versions, when stratified by department.

This higher variation based on training data makes sense given the architecture of fine-tuned ModernBERT. Compared to Llama-3.1-8B, ModernBERT has 1/16^th^ the parameters, which allow for faster fitting to training data during fine-tuning. As such, the need for in-task data (i.e. Mix and NCH-Sleep) is critical to orient the model to the task as it cannot fall back on as much pre-trained parameterization for generalizable performance. Likewise, the increased amount training data in Mix over NCH-Sleep likely helps the model orient itself towards the structure of the task. The Mix trained model is likely able to outperform our best fine-tuned Llama-3.1-8B because its training signal can more dramatically adjust a higher percentage of the parameters in ModernBERT than in the bigger Llama-3.1-8B. In other words, Llama-3.1-8B’s size gives it a inertia that both limits how poor it can do due to generalizability, and how well it can do due to the training signal being more diffused over all of its parameters.

These results suggest that if fine-tuning data is available, constructing an encoder-only classifier is likely more effective for MSH classification than fine-tuning an LLM.

## Experiment 4: Understanding Bias

Having a comprehensive understanding of model bias is critical for any system that is oriented towards clinical application. To evaluate the effects of bias in our best performing systems, we stratify performance by demographics, department, and class. This evaluation will indicate the limitations of these systems as they interact with various facets of real-world application.

### Methods

We evaluate the best performing configuration from experiment 1, 2, and 3, all of which include NCH-Sleep training data. We additionally compare the performance of each configuration when using only MIMIC training data. This additional comparison allows us to see if there is a shift in bias when including our internal data, indicating the presence of stronger or weaker bias signals in NCH-Sleep.

#### Demographics Stratification

It is possible there are disparities in data representation and model performance correlated with population demographics. We evaluate our best models stratified by gender and stratified by race.

#### Department Stratification

Departments vary in how they present author clinical notes, especially due to different norms and EHR templates in use. We evaluate our best models stratified by gender and stratified by department.

#### Performance by Class

Certain MSH classes may be more difficult to classify than others. This may especially shift depending what training data the model has seen. We evaluate how our best model, namely ModernBERT, performs over each class both with and without NCH-Sleep training data. We focus on ModernBERT for this comparison as the ROC curves allow for useful insights into model performance on classes and we find generally these results hold for other models, albeit to a varying magnitude. Additionally, we explore the average span length identified by our expert annotators as positive indicators for each class. It is possible longer span lengths will correlate with increased class representation complexity and therefore worse classification performance.

### Results

#### Performance by Gender

Our results show notable gender disparities in model performance, particularly with discriminative classifiers trained on NCH-Sleep data, achieving substantially higher F1 scores for female-associated notes (0.509) compared to males (0.357). This gap suggests that clinical notes for females contain more explicit or consistent mentions of sleep-related concepts. Conversely, notes for male patients may exhibit greater variability or ambiguity, posing challenges for accurate classification.

#### Performance by Race

We observed consistent racial disparities across models, with substantially better performance for White patients (F1 ∼0.40–0.50) relative to Latino and African American groups (F1 ∼0.25–0.35). The improved classification results for White patients likely reflect higher representational alignment between training data and linguistic patterns commonly documented for these patients. Lower performance for minority groups highlights critical gaps due to linguistic heterogeneity or underrepresentation in clinical data.

#### Performance by Department

Departmental stratification reveals substantial variability, with Behavioral Health notes consistently yielding higher performance (F1 ∼0.45–0.49), likely due to structured and standardized sleep-related documentation practices. In contrast, Primary Care exhibits lower and more variable performance (F1 ∼0.27–0.32), potentially reflecting diverse and less standardized note-taking styles. Performance within smaller departments like School-Based care fluctuates dramatically, emphasizing the impact of training data size and specificity on model robustness.

#### Performance by Class

In Figure 1 we present the ROC curves, and associated AUCs, for our best predictor, ModernBERT trained on Mix, as well as for ModernBERT trained on MIMIC. These curves are presented as an overall performance as well as stratified by MSH class.

**Figure 1:**
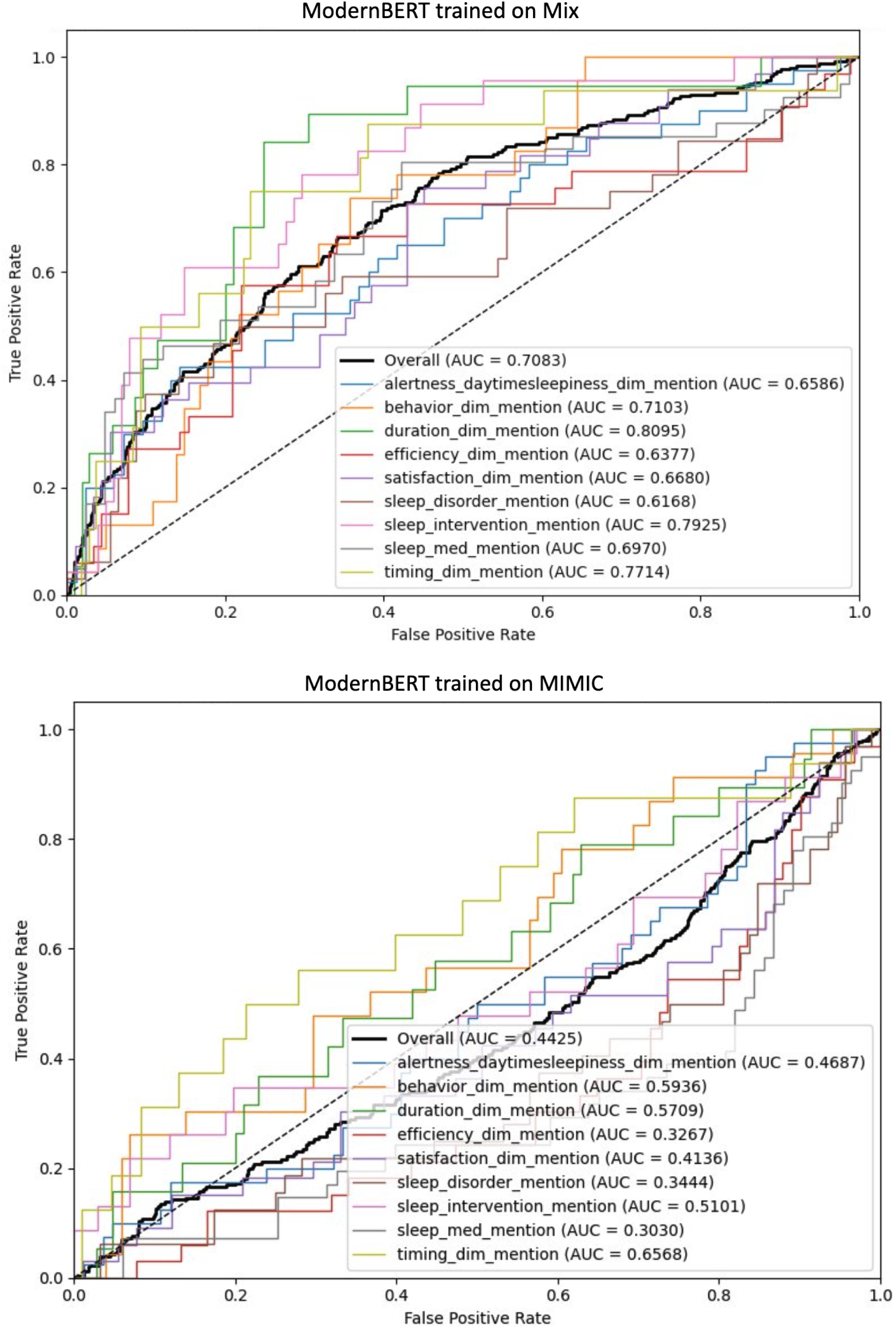
Performance of ModernBERT on NCH-Sleep test, trained on MIX and MIMIC, stratified by MSH Class. ROC curve and AUC ROC presented.

We see in ModernBERT trained on MIMIC, the performance is low for nearly all classes except for sleep timing, behavior, and duration. This implies that these classes are able to build some quantity of a representation using solely MIMIC data. Meanwhile, in ModernBERT trained on Mix, we see these same classes are uplifted while every other class is boosted dramatically. This shows that our NCH-Sleep dataset helps to provide critical information for prediction of the other 6 MSH classes, while also building a better representation over sleep timing, behavior, and duration.

Overall, we see sleep disorder, sleep efficiency, and daytime sleepiness are the hardest behaviors to capture with our classifier. When comparing the average sequence lengths of the indicative spans of these classes, presented in Table 11, we can potentially posit that daytime sleepiness, and especially sleep efficiency, may be harder to classify due to their longer representations in the text. This relationship is tenuous however since sleep disorder has one of the shortest average span lengths and performs poorly while sleep intervention, with the longest average span length, is classified with high accuracy. Likely there are other properties of these classes that indicate difficult representations, perhaps due to irregular lexical choice across notes. We leave this investigation for future work.

**Table 11:** Average span length and standard deviation (characters) per MSH class.

| Class | Average Span Length (std) |
| --- | --- |
| Timing | 24.05 (11.71) |
| Sleep Disorder | 20.74 (16.85) |
| Alertness and Daytime Sleepiness | 25.01 (17.22) |
| Behavior | 38.98 (22.39) |
| Efficiency | 32.49 (18.20) |
| Sleep Medication | 17.25 (17.09) |
| Satisfaction | 22.71 (12.15) |
| Duration | 22.59 (10.75) |
| Sleep Intervention | 38.65 (29.98) |

## Experiment 5: Latency

In clinical application, runtime of a classification system is critical, especially in a screening use-case. If a model takes too much time to run on each note, it is unlikely to be feasibly deployed over an entire healthcare system reviewing all internal notes. A major motivation for creating an AI based classification system is to alleviated the fractured documentation of sleep mentions within clinical practice, making this scalability crucial. Likewise, improved scalability promotes improved transferability into other hospital systems.

### Methods

For each of the following methods we report the average time to provide a prediction for all 100 clinical notes in the NCH-Sleep test set.

#### Dynamic Few-Shot

In this circumstance, we include the latency of encoding the inference example and dynamically gathering few-shot examples for prompt construction. This is paired with the actual model generation time.

#### Fine-tuned LLM

In this circumstance only the latency of the model generation is reported.

#### Fine-tuned Classifier

In this circumstance, only the latency of the model prediction is reported.

### Results

We present our runtime latency results in Table 12. In general, we find a magnitude of difference between the best model of each experiment. Prompting takes the longest, likely due to the need for dynamic few-shot exemplar retrieval using a separate encoder. Fine-tuned Llama-3.1-8B is about 25x faster, showing the large gains to be made when retrieving few-shot exemplars is not a necessary part of inference. Finally, the ModernBERT classifier is even 10x faster than fine-tuned Llama-3.1-8B, and therefore about 250x faster than prompted Llama-3.1-8B. This speed up for ModernBERT is to be expected both because it is a lighter weight model that can directly compute faster, and because it can handle the entire clinical note in its context window at once instead of needing to using a sliding window method.

**Table 12:** Runtime of top performing model from each experiment. Evaluated over 100 NCH-Sleep test notes.

| Model | Average Time to Process Note (Sec) |
| --- | --- |
| Prompted Llama-3.1-8B, 10 Shots, 2000 Window | 18.858 |
| Fine-Tuned Llama-3.1-8B, 8000 window | 0.762 |
| Fine-Tuned ModernBERT Classifier | 0.076 |

These results further show that a fine-tuned encoder-based classifier like ModernBERT can be better suited for specific clinical information extraction tasks than LLMs. This point is especially notable in a live note screening use case where a performative system is deployed at every level of hospital infrastructure to consolidate MSH and provide a comprehensive representation and handling of sleep data. Not only would an LLM system potentially be too slow for this screening task, it performs less accurately when provided with only a small, annotated dataset, and requires more specialty computing equipment like state-of-the-art GPUs.

## Conclusion

Overall, our findings suggest the best system for MSH classification is either a fine-tuned encoder-based classifier with a combination of related-data and task specific data or a prompted LLM with as many few-shot exemplars as possible. The decision should be made based on if training data is available or not. Fine-tuning an LLM is a less effective use of training data than finetuning a classifier but prompting an LLM will do when no data is available at all.

### Future Direction

#### Other Model Architectures

In this work we primarily compare transformer architectures with two decoder-only LLMs and one encoder-only LLM. Several other model architectures have shown promise in recent works, including state-space-models (SSMs) such as Mamba, and Mixture-of-Expert (MoE) models such as Mixtral [12, 13]. SSMs deviate from standard transformer architecture with claims of especially fast runtime, providing a potential alternative to our encoder-only classifier implemented by ModernBERT, our fastest solution according to latency evaluation [12]. Meanwhile, MoE systems tout improved model training on specific tasks, due to its expert-gating mechanism, and improved runtime relative to parameter size, providing an alternative to our LLM approaches [13].

#### Future Direction: Chain-of-thought Prompting

Contemporary prompting techniques promote reasoning over a task using LLM in-context reasoning capabilities. This is generally employed by having the model generate intermediary thoughts in a sequence leading up to the solution, referred to as Chain-of-Thought (CoT). This method may be effective for our complex MSH classification task but does not have a straightforward implementation. One possible option is to have the LLM first identify the spans, as per our annotations, by searching for relevant words for the provided class before then making a prediction. Our preliminary efforts in this direction, however, ran into issues as many times the LLM would wander off the span-extraction task and start hallucinating. The classification generated from a hallucinated CoT process, if it was generated at all, was often incorrect. We found in 20% of cases no final classification was provided and often performance was poor when it was. It is possible that newer LLMs may perform better on this task on our dataset due to their improved efficiency.

## Data Availability

The data for this study is not available

## Appendix

## A1) Validating Dynamic Few-Shot Prompting

We initially validate our prompting methods using GPT-4o-mini and MIMIC data [8]. The use of GPT-4o-mini allows for rapid testing due to API calls being faster than local compute. Additionally, GPT-4o-mini performance is well verified and considered high overall, allowing isolation of independent variables related to the prompt [8]. We use the MIMIC dataset (combination of MDACE and SDOH as mentioned in Table 4) because the notes are deidentified and can be sent to an API. Additionally, MIMIC has been tested in other works and provides a benchmark of replicability.

### Method

We evaluate GPT-4o-mini on a randomly selected sample of 1000 examples from the MIMIC dataset with another 1000 held out as our training set. We perform dynamic few-shot prompting, as described in Experiment 1, using openAI’s text-embedding-3-large as our encoder. We encode our training set and gather *n* few-shot examples using cosine distance from the test-example embedding. GPT-4o-mini has a large context window that allows us to avoid windowing.

### Results

In this stage we also experimented with other prompts, iteratively improving until we arrived upon the prompt presented in experiment 1. One of the major improvements came from asking the model to classify “true” / “false” rather than 1 / 0. The numbers are only reported for the final “true” / “false” version.

Table A1 presents our preliminary results on validating few-shot prompting using MIMIC data and gpt-4o-mini. We see markedly high performance

**Table A1.** Evaluation of dynamic few-shot prompting using gpt-4o-mini and notes from our MIMIC training set.

| #-Shots | F1 |
| --- | --- |
| 1 | 0.90 |
| 3 | 0.90 |
| 2 | 0.89 |
| 5 | 0.89 |
| 7 | 0.88 |
| 0 | 0.88 |
| 10 | 0.88 |

## A2) Complete Prompting Results

**Table A2:** Complete Prompting Results from Experiment 1.

| model | shots | window | precision | recall | f1 | Token prob AUC |
| --- | --- | --- | --- | --- | --- | --- |
| Llama-3.1-8B | 5 | 1000 | 0.2307 | 0.9471 | 0.371 | 0.6743 |
| Llama-3.1-8B | 7 | 1000 | 0.2376 | 0.9365 | 0.379 | 0.6631 |
| Llama-3.1-8B | 10 | 1000 | 0.2347 | 0.9153 | 0.3737 | 0.6471 |
| Llama-3.1-8B | 5 | 2000 | 0.2408 | 0.8307 | 0.3734 | 0.6444 |
| Llama-3.1-8B | 10 | 2000 | 0.25 | 0.8307 | 0.3843 | 0.636 |
| Llama-3.1-8B | 7 | 2000 | 0.2437 | 0.8148 | 0.3752 | 0.6356 |
| Llama-3.1-8B | 3 | 1000 | 0.2196 | 0.9947 | 0.3598 | 0.6298 |
| Llama-3.1-8B | 7 | 2000 | 0.2105 | 1 | 0.3477 | 0.6288 |
| Llama-3.1-8B | 7 | 8000 | 0.2102 | 1 | 0.3474 | 0.6199 |
| Llama-3.1-8B | 5 | 2000 | 0.2108 | 0.9947 | 0.3478 | 0.6159 |
| Llama-3.1-8B | 3 | 2000 | 0.2354 | 0.8519 | 0.3688 | 0.6133 |
| Llama-3.1-8B | 10 | 2000 | 0.21 | 1 | 0.3471 | 0.6076 |
| Llama-3.1-8B | 10 | 8000 | 0.2102 | 1 | 0.3474 | 0.6035 |
| BioMistral-7B | 10 | 8000 | 0.2606 | 0.6898 | 0.3783 | 0.5953 |
| Llama-3.1-8B | 10 | 4000 | 0.2514 | 0.7354 | 0.3747 | 0.5924 |
| Llama-3.1-8B | 7 | 4000 | 0.2509 | 0.7354 | 0.3742 | 0.5853 |
| Llama-3.1-8B | 0 | 1000 | 0.21 | 1 | 0.3471 | 0.5849 |
| BioMistral-7B | 3 | 6000 | 0.2484 | 0.8307 | 0.3825 | 0.5841 |
| Llama-3.1-8B | 5 | 8000 | 0.2136 | 0.9947 | 0.3517 | 0.5833 |
| BioMistral-7B | 7 | 6000 | 0.2609 | 0.6984 | 0.3799 | 0.5805 |
| Llama-3.1-8B | 7 | 1000 | 0.2102 | 1 | 0.3474 | 0.5802 |
| BioMistral-7B | 7 | 2000 | 0.2187 | 0.9894 | 0.3582 | 0.5797 |
| Llama-3.1-8B | 7 | 6000 | 0.2098 | 0.9947 | 0.3465 | 0.5793 |
| Llama-3.1-8B | 3 | 1000 | 0.2112 | 1 | 0.3487 | 0.5781 |
| BioMistral-7B | 10 | 6000 | 0.2528 | 0.7143 | 0.3734 | 0.5778 |
| Llama-3.1-8B | 7 | 10000 | 0.2105 | 1 | 0.3477 | 0.5755 |
| BioMistral-7B | 7 | 8000 | 0.2209 | 0.8148 | 0.3476 | 0.572 |
| Llama-3.1-8B | 10 | 6000 | 0.25 | 0.7037 | 0.3689 | 0.5713 |
| Llama-3.1-8B | 5 | 6000 | 0.2111 | 0.9894 | 0.3479 | 0.5713 |
| Llama-3.1-8B | 10 | 1000 | 0.21 | 1 | 0.3471 | 0.5703 |
| BioMistral-7B | 5 | 8000 | 0.2249 | 0.8503 | 0.3557 | 0.5701 |
| Llama-3.1-8B | 5 | 10000 | 0.2144 | 0.9894 | 0.3525 | 0.5697 |
| Llama-3.1-8B | 7 | 10000 | 0.2436 | 0.7513 | 0.3679 | 0.5695 |
| BioMistral-7B | 10 | 4000 | 0.2279 | 0.8995 | 0.3636 | 0.5682 |
| BioMistral-7B | 5 | 6000 | 0.2222 | 0.8783 | 0.3547 | 0.5682 |
| BioMistral-7B | 3 | 2000 | 0.2143 | 0.9683 | 0.3509 | 0.5682 |
| BioMistral-7B | 0 | 6000 | 0.2292 | 0.9048 | 0.3658 | 0.5674 |
| Llama-3.1-8B | 5 | 1000 | 0.21 | 1 | 0.3471 | 0.5673 |
| BioMistral-7B | 0 | 1000 | 0.2105 | 1 | 0.3477 | 0.5663 |
| BioMistral-7B | 0 | 10000 | 0.2284 | 0.8389 | 0.3591 | 0.5662 |
| BioMistral-7B | 3 | 10000 | 0.2339 | 0.7672 | 0.3585 | 0.5651 |
| Llama-3.1-8B | 10 | 10000 | 0.2107 | 1 | 0.3481 | 0.5649 |
| BioMistral-7B | 3 | 4000 | 0.2228 | 0.8995 | 0.3571 | 0.5645 |
| BioMistral-7B | 10 | 6000 | 0.2272 | 0.8836 | 0.3615 | 0.5644 |
| BioMistral-7B | 7 | 4000 | 0.2119 | 0.8307 | 0.3376 | 0.5644 |
| Llama-3.1-8B | 3 | 2000 | 0.2105 | 0.9947 | 0.3475 | 0.5639 |
| Llama-3.1-8B | 3 | 10000 | 0.2154 | 0.9894 | 0.3538 | 0.5637 |
| BioMistral-7B | 7 | 4000 | 0.2299 | 0.6898 | 0.3449 | 0.5632 |
| BioMistral-7B | 0 | 2000 | 0.2121 | 0.9841 | 0.349 | 0.5631 |
| BioMistral-7B | 3 | 2000 | 0.2166 | 0.9788 | 0.3547 | 0.562 |
| BioMistral-7B | 3 | 8000 | 0.2153 | 0.8245 | 0.3414 | 0.562 |
| BioMistral-7B | 5 | 2000 | 0.2153 | 0.9841 | 0.3533 | 0.5615 |
| BioMistral-7B | 0 | 8000 | 0.2314 | 0.8777 | 0.3663 | 0.5602 |
| Llama-3.1-8B | 10 | 6000 | 0.2102 | 1 | 0.3474 | 0.5598 |
| BioMistral-7B | 5 | 6000 | 0.2418 | 0.7074 | 0.3604 | 0.5588 |
| BioMistral-7B | 0 | 4000 | 0.2186 | 0.9471 | 0.3552 | 0.5556 |
| Llama-3.1-8B | 5 | 4000 | 0.2473 | 0.7302 | 0.3695 | 0.555 |
| Llama-3.1-8B | 10 | 10000 | 0.2217 | 0.7249 | 0.3395 | 0.5537 |
| BioMistral-7B | 7 | 6000 | 0.2144 | 0.8201 | 0.3399 | 0.5533 |
| BioMistral-7B | 7 | 10000 | 0.2204 | 0.788 | 0.3444 | 0.5525 |
| BioMistral-7B | 10 | 4000 | 0.2462 | 0.6809 | 0.3616 | 0.5521 |
| BioMistral-7B | 7 | 10000 | 0.2635 | 0.5989 | 0.366 | 0.5516 |
| BioMistral-7B | 5 | 2000 | 0.2217 | 0.9735 | 0.3611 | 0.5508 |
| BioMistral-7B | 7 | 2000 | 0.236 | 0.9577 | 0.3787 | 0.5497 |
| BioMistral-7B | 5 | 10000 | 0.2622 | 0.6596 | 0.3752 | 0.5485 |
| BioMistral-7B | 7 | 1000 | 0.2112 | 0.9947 | 0.3485 | 0.5482 |
| Llama-3.1-8B | 5 | 6000 | 0.2486 | 0.6878 | 0.3652 | 0.547 |
| BioMistral-7B | 10 | 10000 | 0.2557 | 0.6188 | 0.3619 | 0.5464 |
| BioMistral-7B | 10 | 2000 | 0.2436 | 0.8564 | 0.3793 | 0.5459 |
| BioMistral-7B | 3 | 6000 | 0.2332 | 0.8254 | 0.3636 | 0.5453 |
| Llama-3.1-8B | 5 | 10000 | 0.2403 | 0.7566 | 0.3648 | 0.5444 |
| BioMistral-7B | 5 | 1000 | 0.2124 | 1 | 0.3503 | 0.5421 |
| BioMistral-7B | 10 | 2000 | 0.22 | 0.9788 | 0.3592 | 0.5415 |
| Llama-3.1-8B | 10 | 8000 | 0.2342 | 0.75 | 0.357 | 0.541 |
| Llama-3.1-8B | 7 | 6000 | 0.2533 | 0.7037 | 0.3725 | 0.5406 |
| BioMistral-7B | 7 | 8000 | 0.2439 | 0.6898 | 0.3603 | 0.5396 |
| BioMistral-7B | 5 | 10000 | 0.2212 | 0.7872 | 0.3454 | 0.5381 |
| Llama-3.1-8B | 3 | 6000 | 0.2139 | 0.9788 | 0.351 | 0.5365 |
| Llama-3.1-8B | 7 | 8000 | 0.2491 | 0.7513 | 0.3742 | 0.5363 |
| BioMistral-7B | 3 | 4000 | 0.2331 | 0.9101 | 0.3711 | 0.5363 |
| Llama-3.1-8B | 3 | 8000 | 0.2163 | 0.9841 | 0.3546 | 0.5349 |
| Llama-3.1-8B | 10 | 4000 | 0.2102 | 1 | 0.3474 | 0.5341 |
| BioMistral-7B | 5 | 4000 | 0.2246 | 0.7831 | 0.3491 | 0.5332 |
| BioMistral-7B | 10 | 1000 | 0.2152 | 0.9894 | 0.3535 | 0.5318 |
| BioMistral-7B | 5 | 4000 | 0.2257 | 0.9101 | 0.3617 | 0.5309 |
| BioMistral-7B | 3 | 10000 | 0.2621 | 0.6649 | 0.3759 | 0.5305 |
| BioMistral-7B | 7 | 1000 | 0.2112 | 0.9947 | 0.3485 | 0.5303 |
| BioMistral-7B | 5 | 8000 | 0.2308 | 0.7021 | 0.3474 | 0.53 |
| Llama-3.1-8B | 0 | 2000 | 0.21 | 1 | 0.3471 | 0.5297 |
| BioMistral-7B | 3 | 8000 | 0.228 | 0.7287 | 0.3473 | 0.5284 |
| Llama-3.1-8B | 3 | 4000 | 0.2141 | 0.9947 | 0.3524 | 0.5281 |
| BioMistral-7B | 3 | 1000 | 0.2112 | 1 | 0.3487 | 0.5272 |
| Llama-3.1-8B | 0 | 6000 | 0.21 | 1 | 0.3471 | 0.5226 |
| Llama-3.1-8B | 7 | 4000 | 0.2114 | 1 | 0.349 | 0.5217 |
| BioMistral-7B | 10 | 10000 | 0.209 | 0.7636 | 0.3281 | 0.5216 |
| Llama-3.1-8B | 3 | 4000 | 0.2496 | 0.8042 | 0.381 | 0.5209 |
| BioMistral-7B | 10 | 1000 | 0.2121 | 1 | 0.35 | 0.5199 |
| BioMistral-7B | 5 | 1000 | 0.211 | 0.9947 | 0.3481 | 0.5189 |
| BioMistral-7B | 10 | 8000 | 0.2267 | 0.8389 | 0.357 | 0.518 |
| Llama-3.1-8B | 3 | 6000 | 0.2416 | 0.7196 | 0.3617 | 0.5139 |
| Llama-3.1-8B | 5 | 4000 | 0.2109 | 1 | 0.3484 | 0.5077 |
| BioMistral-7B | 3 | 1000 | 0.212 | 0.9947 | 0.3494 | 0.5008 |
| Llama-3.1-8B | 5 | 8000 | 0.2482 | 0.7407 | 0.3718 | 0.4958 |
| Llama-3.1-8B | 3 | 10000 | 0.2373 | 0.7937 | 0.3654 | 0.4751 |
| Llama-3.1-8B | 0 | 8000 | 0.21 | 1 | 0.3471 | 0.4709 |
| Llama-3.1-8B | 0 | 4000 | 0.21 | 1 | 0.3471 | 0.4581 |
| Llama-3.1-8B | 0 | 10000 | 0.21 | 1 | 0.3471 | 0.4483 |
| Llama-3.1-8B | 3 | 8000 | 0.2508 | 0.7937 | 0.3812 | 0.4463 |

## Notes

### Competing Interest Statement

The authors have declared no competing interest.

### Funding Statement

MD was supported by the National Heart, Lung, Blood Institute (1K01HL169493-1; Principal Investigator: MD)

### Author Declarations

This study was approved by Nationwide Children's Hospital's internal IRB STUDY00002624.

